# Comparative validation of the BOADICEA and Tyrer-Cuzick breast cancer risk models incorporating classical risk factors and polygenic risk in a population-based prospective cohort

**DOI:** 10.1101/2020.04.27.20081265

**Authors:** Parichoy Pal Choudhury, Mark N. Brook, Amber N. Wilcox, Andrew Lee, Charlotta Mulder, Penny Coulson, Minouk J. Schoemaker, Michael E. Jones, Anthony J. Swerdlow, Nilanjan Chatterjee, Antonis C. Antoniou, Montserrat Garcia-Closas

**Author notes:** Corresponding author: Dr. Montserrat Garcia-Closas, Division of Cancer Epidemiology and Genetics, National Cancer Institute, National Institutes of Health, 9609 Medical Center Drive 7E-342, Rockville, MD 20850, USA.

## Abstract

**Purpose:** The Breast and Ovarian Analysis of Disease Incidence and Carrier Estimation Algorithm (BOADICEA) and the Tyrer-Cuzick breast cancer models have recently been extended to include polygenic risk scores (PRS). In addition, BOADICEA has also been extended to include reproductive and lifestyle factors, which were already part of Tyrer-Cuzick model. We conducted a comparative validation of the extended models including a recently developed 313-variant PRS in a population-based prospective cohort.

**Methods:** We used data from a nested case-control sample of 1,337 women of European ancestry (619 incident breast cancer cases) aged 23-75 years from the Generations Study. Models were evaluated for calibration of five-year absolute risk and risk discrimination.

**Results:** The extended BOADICEA model with risk factors and PRS was well calibrated across risk deciles: expected-to-observed ratio (E/O) at the highest risk decile = 0.97 (95% CI = 0.51 to 1.86) for women younger than 50 years and 1.09 (0.66 to 1.80) for women 50 years or older. Adding risk factors and PRS to the BOADICEA model improved discrimination modestly in younger women (Area Under the Curve (AUC): 69.7% vs. 69.1%) and substantially in older women (AUC: 64.6% vs. 56.8%). The Tyrer-Cuzick model with PRS had similar discrimination as the extended BOADICEA model for both age groups; but showed evidence of overestimation at the highest risk decile: E/O=1.54 (0.81 to 2.92) for younger and 1.73 (1.03 to 2.90) for older women.

**Conclusion:** The extended BOADICEA model identified women in a European ancestry population at elevated breast cancer risk more accurately than the Tyrer-Cuzick model with PRS. These analyses can inform choice of risk models for risk stratified breast cancer prevention for women of European ancestry.

## Introduction

The Breast and Ovarian Analysis of Disease and Carrier Estimation Algorithm (BOADICEA) breast cancer model was originally developed to predict breast cancer risk for women using pedigree-level family history information and genetic testing results on rare pathogenic variants in high and moderate risk genes (1–4). This model has been updated (version 5.0) to include reproductive and lifestyle factors and the recently developed polygenic risk score (PRS) based on 313 common germline variants (5) for applications in both general and high-risk populations (6). The Tyrer-Cuzick or International Breast Intervention Study (IBIS) model (7), commonly used in clinical and research settings, also includes extensive family history and comprehensive risk factor information, and has been updated to include information on PRS. We recently evaluated the performance of the Tyrer-Cuzick model (version 8.0) without PRS in a prospective cohort of women of European ancestry in the general population (8). Here, we perform a comparative validation of the extended BOADICEA and Tyrer-Cuzick models incorporating the 313-variant PRS in the same prospective cohort.

The original BOADICEA and Tyrer-Cuzick models are considered in clinical guidelines (9,10) for management of women with a family history of breast cancer, and have been implemented in user-friendly risk assessment tools (BOADICEA: https://canrisk.org; IBIS: https://ibis.ikonopedia.com). Given their widespread use, comparative prospective validation of these models and their extensions is critical to assess their ability to accurately identify women at different risk levels for risk-stratified screening, surveillance or prevention strategies.

We report results from a prospective comparative validation of the extended BOADICEA model (v.5) with risk factors and 313-variant PRS and Tyrer-Cuzick model (v.8) with the same PRS in the Generations Study, a population-based cohort study of UK women (11).

## Methods

Data were used from a nested case-control sample within the Generations Study (2003–2012), a prospective cohort of over 113,000 UK women aged 16–102 years, details are elsewhere (8,11). The comparative validation analyses were based on 1,337 women aged 23–75 years, including 619 incident breast cancer patients within five-years from study recruitment, with information on the PRS and the risk factors used in both the BOADICEA (v.5) and Tyrer-Cuzick (v.8) models.

To update the original BOADICEA model, the relative risks for the risk factors and PRS were derived using the literature-based approach (5,8); further details are given in Lee *et al*. (6). In this model, the family history association, described by a residual polygenic component, was adjusted to account for the PRS explaining ~20% of the breast cancer familial aggregation. The PRS was added to the Tyrer-Cuzick model (v.8) using the approach described in Brentnall *et al*. (12), where the associations of family history and PRS were unadjusted and assumed to be multiplicative on the risk of developing breast cancer. The comparative validation analyses were performed using the standardized model calibration and discrimination methods implemented in the Individualized Coherent Absolute Risk Estimator (iCARE) tool (13) (details in supplement). Briefly, model calibration was assessed in terms of relative and absolute risk by comparing the observed and expected quantities, overall and within risk categories. The Area Under the Curve (AUC) was estimated to assess model discrimination.

## Results

For women younger than 50 years, the original and extended BOADICEA models (with PRS and with PRS and risk factors) showed good calibration of relative and absolute risk (Figure 1). At the highest decile of predicted five-year absolute risk, the extended model with PRS and risk factors showed better calibration than both the original model and the extended model with PRS only, with a ratio of expected to observed number of cases (E/O) of 0.97 [95% confidence interval (CI): 0.51 to 1.86], 0.83 (0.44 to 1.56), 0.85 (0.44 to 1.63), respectively. Adding PRS and risk factors led to modest improvement in AUC from 69.1% (63.5% to 74.6%) to 69.7% (64.1% to 75.2%). Incorporating risk factors did not improve the discrimination of the original model (data not shown) or the extended model with PRS (Figure 1). The Tyrer-Cuzick model with PRS had similar discrimination [AUC: 69.4% (63.8% to 75.0%)] to the extended BOADICEA model with PRS and risk factors but showed evidence of overestimation at the highest risk decile [E/O:1.54 (0.81 to 2.92)].

**Figure 1:**
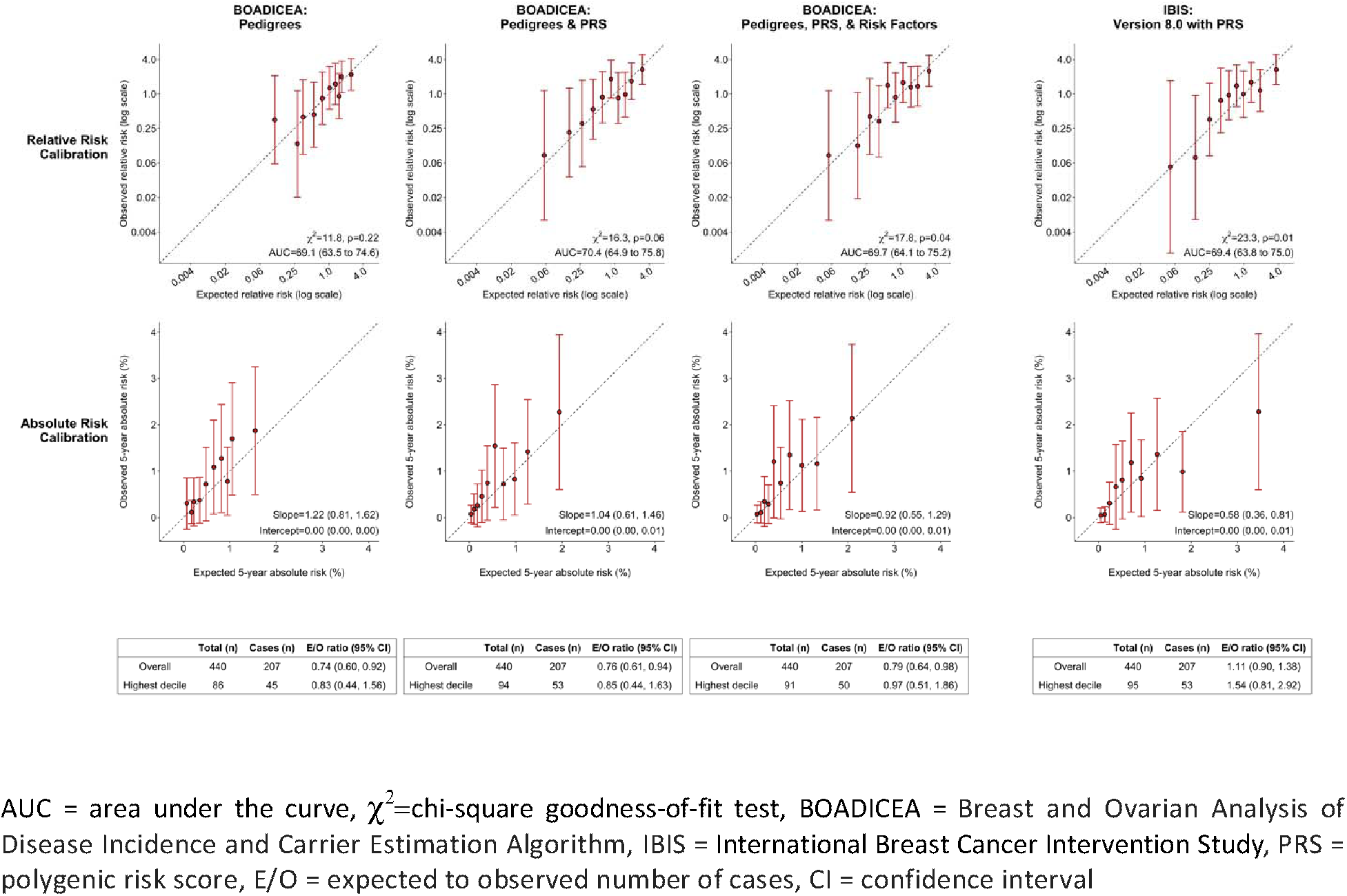
Calibration and discrimination of five-year risk predictions of breast cancer for women younger than 50 years in the nested case-control sample of the Generations Study cohort with risk categories based on deciles of predicted five-year absolute risk. Validation results are shown for the original BOADICEA model that incorporates pedigree level family history information, its two extensions: (i) incorporating the recently developed PRS based on 313 common germline variants to the original model and (ii) incorporating the 313-variant PRS and reproductive and lifestyle factors to the original model, and the IBIS (Version 8.0) model after including the 313-variant PRS. Estimates and 95% Cl of the calibration slope and intercept are reported based on a linear regression of the decile-specific observed proportion of cases within five years and the average of the predicted five-year absolute risk.

The original and extended BOADICEA models also showed good calibration of relative and absolute risk for women 50 years or older (Figure 2), in particular for women at the highest risk decile [E/O: 0.95 (0.56 to 1.62) for the original model, 1.07 (0.63 to 1.82) for the extended model with PRS, 1.09 (0.66 to 1.80) for the extended model with PRS and risk factors]. For this age group, incorporating PRS and risk factors led to substantial improvements in AUC from 56.8% (52.9% to 60.6%) to 64.6% (60.9% to 68.2%). Adding risk factors substantially improved the risk discrimination of the original model (data not shown) and the extended model with PRS (Figure 2). The Tyrer-Cuzick model with PRS [AUC: 63.9% (60.2% to 67.6%)] had risk discrimination comparable to the extended BOADICEA model with PRS and risk factors; however, the former substantially overestimated risk for women at the highest risk decile [E/O: 1.73 (1.03 to 2.90)].

**Figure 2:**
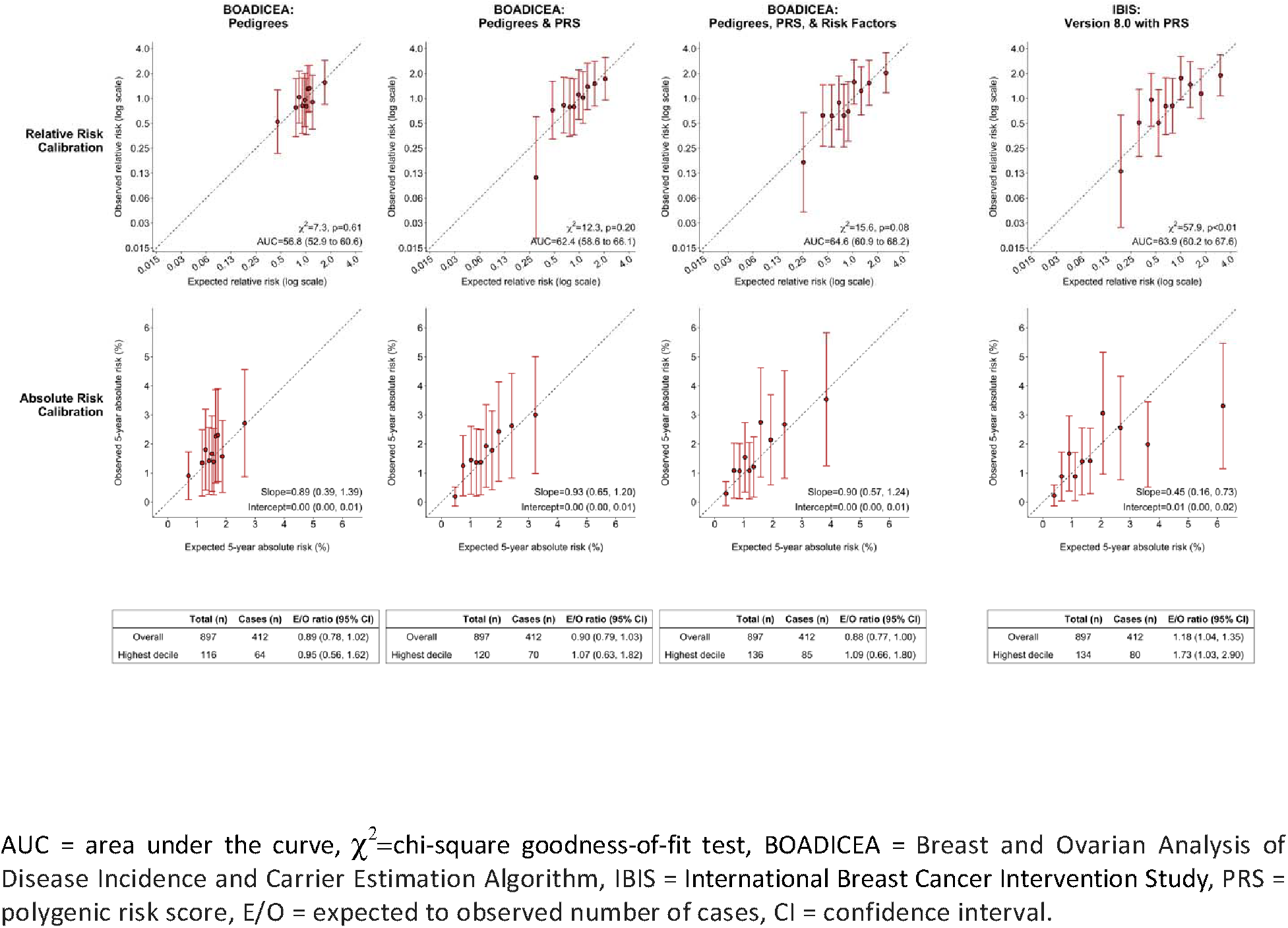
Calibration and discrimination of five-year risk predictions of breast cancer for women aged 50 years or older in the nested case-control sample of the Generations Study cohort with risk categories based on deciles of predicted five-year absolute risk. Validation results are shown for the original BOADICEA model that incorporate pedigree level family history information, its two extensions: (i) incorporating the recently developed PRS based on 313 common germline variants to the original model and (ii) incorporating the 313-variant PRS and reproductive and lifestyle factors to the original model, and the IBIS model (Version 8.0) after including the 313-variant PRS. Estimates and 95% Cl of the calibration slope and intercept are reported based on a linear regression of the decile-specific observed proportion of cases within five years and the average of the predicted five-year absolute risk.

## Discussion

Our study shows that the extended BOADICEA model, which incorporated reproductive and lifestyle factors and a 313-SNP PRS to the familial aggregation information, predicted five-year absolute risk of breast cancer more accurately than the Tyrer-Cuzick model with the same PRS, for women at the highest risk decile in the Generations study, a UK-based prospective cohort.

Previous studies in populations of women of European ancestry provided evidence of overestimation of absolute risk obtained from the Tyrer-Cuzick model without PRS for women in the highest risk decile (8,14). Two recent studies that incorporated PRSs with fewer genetic variants to this model, showed good calibration in terms of relative risk, but did not evaluate absolute risk calibration (12,15). Our results showed that incorporation of the 313-SNP PRS worsened the absolute risk calibration for women at the high-risk deciles (Supplementary Figure 1), possibly due to not attenuating the contribution of family history association to account for the substantial familial aggregation explained by the PRS. This can lead to inflated breast cancer risks, particularly for women with breast cancer family history who are more prevalent in high-risk deciles.

Strengths of the current analyses include the use of the Generations Study, a relatively recent population-based cohort with a wide range of ages of participating women and the comparison of two widely used risk prediction tools. Moreover, model calibration was assessed both overall and within risk categories, in particular for women at the extremes of risk for whom prevention and screening are most relevant. While the current study provides some evidence of accurate risk predictions from the CanRisk tool for the UK general population, further evaluation of this tool in both general and high-risk populations is needed before widespread clinical applications.

## Conclusion

In this study, the extended BOADICEA model with PRS and risk factors identified women of European ancestry at elevated five-year risk of breast cancer more accurately than the Tyrer-Cuzick model with PRS. These analyses can inform the choice of risk models for developing risk-stratified breast cancer prevention and screening strategies for women of European ancestry. Further research on risk model building and validation is needed for developing such strategies for women of non-European ancestry.

## Data Availability

N/A

## Acknowledgement

This work was supported by Cancer Research UK grants C12292/A20861 and C1287/A16563; the European Union’s Horizon 2020 research and innovation programme under grant agreement number 633784 (B-CAST). The Generations Study acknowledge Breast Cancer Now and the Institute of Cancer Research for support and funding of the Generations Study, and the study participants, study staff, and the doctors, nurses and other healthcare staff and data providers who have contributed to the study. The Institute of Cancer Research acknowledge National Health Service funding to the National Institute for Health Research Biomedical Research Center, UK. The works of Parichoy Pal Choudhury, Montserrat Garcia-Closas were supported by the Intramural Research Program, Division of Cancer Epidemiology and Genetics, National Cancer Institute, National Institutes of Health, Department of Health and Human Services. The work of Amber N. Wilcox is supported by the University of North Carolina LCCC Cancer Control Education Program predoctoral training grant from the National Cancer Institute (T32-CA057726). The work of Nilanjan Chatterjee was supported by the Patient Centered Outcomes Research Institute (PCORI) Award (ME1602–34530). We would like to thank Adam R. Brentnall and Jack Cuzick for providing the IBIS Risk Evaluator software.

